# Conditionally positive: a qualitative study of public perceptions about using health data for artificial intelligence research

**DOI:** 10.1101/2020.04.25.20079814

**Authors:** Melissa D McCradden, Tasmie Sarker, P Alison Paprica

## Abstract

**Objectives:** Given widespread interest in applying artificial intelligence (AI) to health data to improve patient care and health system efficiency, there is a need to understand the perspectives of the general public regarding the use of health data in AI research.

**Design:** A qualitative study involving six focus groups with members of the public. Participants discussed their views about AI in general, then were asked to share their thoughts about three realistic health AI scenarios. Data were analysed using qualitative description thematic analysis.

**Settings:** Two cities in Ontario, Canada: Sudbury (400 km north of Toronto) and Mississauga, (part of the Greater Toronto Area).

**Participants:** Forty-one purposively sampled members of the public (21M:20F, 25-65 years, median age 40).

**Results:** Participants had low levels of prior knowledge of AI and mixed, mostly negative, perceptions of AI in general. Most endorsed AI as a tool for the analysis of health data when there is strong potential for public benefit, providing that concerns about privacy, consent, and commercial motives were addressed. Inductive thematic analysis identified AI-specific hopes (e.g., potential for faster and more accurate analyses, ability to use more data), fears (e.g., loss of human touch, skill depreciation from over-reliance on machines) and conditions (e.g., human verification of computer-aided decisions, transparency). There were mixed views about whether consent is required for health data research, with most participants wanting to know if, how and by whom their data were used. Though it was not an objective of the study, realistic health AI scenarios were found to have an educational effect.

**Conclusions:** Notwithstanding concerns and limited knowledge about AI in general, most members of the general public in six focus groups in Ontario, Canada perceived benefits from health AI and conditionally supported the use of health data for AI research.

**STRENGTHS AND LIMITATIONS OF THIS STUDY:** A strength of this study is the analysis of how diverse members of the general public perceive three realistic scenarios in which health data are used for AI research.

The detailed health AI scenarios incorporate points that previous qualitative research has indicated are likely to elicit discussion (e.g., use of health data without express consent, involvement of commercial organisations in health research, inability to guarantee anonymity of genetic data) and may also be useful in future qualitative research studies and for educational purposes.

The findings are likely to be relevant to organisations that are considering making health data available for AI research and development.

Notwithstanding the diverse ethnic and educational backgrounds of participants, overall the sample represents the general (mainstream) population of Ontario and results cannot be interpreted as presenting the views of specific subpopulations and may not be generalisable across Ontario or to other settings.

Given the low level of knowledge about AI in general it is possible that the views of participants would change substantially if they learned and understood more about AI.

**TRANSPARENCY STATEMENT:** P. Alison Paprica affirms that the manuscript is an honest, accurate and transparent account of the study being reported; that no important aspects of the study have been omitted; and that there were no discrepancies from the study as originally approved by the University of Toronto Research Ethics Board.

## INTRODUCTION

Modern artificial intelligence (AI) and its subfield machine learning (ML) offer much promise for deriving impactful knowledge from health data. Several recent articles present summaries of recent health AI and ML achievements, and what the future could look like as more health data become available and are used in AI research and development[1–5]. Given that AI and ML require large amounts of data,[6] public trust in, and support for, using health data for AI/ML will be essential. Many institutions are exploring models for using large representative datasets of health information to create learning healthcare systems[7,8]. Public trust and social licence for such work is essential[8] because, in contrast with clinical studies that have consent-based participation from data subjects, “big data” research is often performed without expressed consent from the data subjects[9]. Previous studies exploring the public attitudes toward data-intensive health research in general, i.e., without an AI/ML focus, found that most members of the mainstream public are supportive provided there are appropriate controls[10–13]. While underscoring the need to address the public’s concerns, studies in Canada, the UK, USA and other jurisdictions suggest that members of the mainstream public view health data as an asset that should be used as long as their concerns related to privacy, commercial motives and other risks are addressed[10–13].

However, we cannot assume that this general but conditional public support for data-intensive health research extends to AI/ML for several reasons. Foremost, research has shown that the members of the general public have low understanding of AI in general, alongside AI-specific hopes and fears including loss of control of AI, ethical concerns, and the potential negative impact of AI on work[14–18]. Secondly, while there is general trend toward support for health AI,[19] there is also recent negative press about large technology companies using health data for AI, including patients suing Google and the University of Chicago Medical Center[20] and the view of the National Data Guardian at the UK’s Department of Health that the sharing of patient data between the Royal Free Hospital of London and Google DeepMind was legally inappropriate[21]. Thirdly, there is decreasing confidence that accepted approaches to de-identification are sufficient to ensure privacy in the face of AI’s capabilities[22].

To date, there has been limited scholarly research on public perceptions of health AI. Most published studies have focused on the views of patients who may not be representative because they stand to benefit from AI applications[16]. Further, most published studies have focused on computer vision health AI applications in radiology and dermatology, which represent only a small fraction of the potential applications of AI in health[23–25]. Additionally, there is a need to understand public perspectives versus patient perspectives, because health AI research may rely on large datasets that include information about people who do not have health conditions and/or do not stand to benefit directly from the research. Accordingly, the objective of this study was to learn more about how members of the general public perceive health data being used for AI research.

## METHODS

### Study Design

Focus groups were conducted using semi-structured discussion guides designed to prompt dialogue among participants (see supplemental materials). Each two-hour focus group had four parts: (i) warm-up exercise and participant views about AI in general, (ii) brief introduction of the Vector Institute for artificial intelligence (Vector) and plain language examples of AI/ML supplied by Vector, (iii) of participant views on realistic but fictional health AI scenarios (see supplemental materials), and (iv) time for questions with a Vector representative (PAP). The three scenarios were presented in varying order across groups per site, and included AI-based Cancer Genetics Test, an AI-based App to Help Older Adults Aging at Home, and an Accessible Health Dataset of Lab Test Results for AI. Participants were asked to make an independent written decision about the acceptability of each health AI scenario before the group discussion began to increase the likelihood that they would state their own initial views versus echo the views of others. The study was approved by the Research Ethics Board of the University of Toronto in Toronto, Ontario, Canada, protocol number 38084.

### Setting

The sessions took place in October 2019 in facilities designed for focus groups with audio-recording capabilities and space for observation (PAP, MDM, TS) behind a one-way mirror. Three focus groups were conducted in northern Ontario (Sudbury, 400 km north of Toronto) and three in the Greater Toronto Area (Mississauga).

### Participants

A total of 41 participants took part in the research (Table 1) – 20 participants in Sudbury, 21 participants in Mississauga. Participants were contacted through Canadian Market Research, drawing from a repository of individuals who had consented to be contacted for research. Purposive sampling was used to identify eight invitees for each focus group that collectively had variation in age, gender, income, education, ethnicity and household size[26]. Of the 48 people approached, one person arrived unwell and was compensated but sent home, and six did choose to attend (reasons not captured). To create an environment in which participants were most likely to be comfortable sharing their views, in each city there was an afternoon focus group with individuals ages 25-34 and mixed incomes, followed by 5:00 pm focus group with people ages 35-65 with lower incomes, and a 7:30 pm focus group with people ages 35-65 and higher incomes. For practical reasons, recruitment for all focus groups occurred at one time. As part of the recruitment process, participants were notified of the purpose of the focus groups, i.e., to learn more about how members of the public perceive the use of health data for AI research. Participants were also informed of the purpose of each focus group, in writing, as part of the process to obtain their written informed consent immediately before each session, and verbally at the start of each focus group. At the end of each session, participants were provided with a cheque for $100 CAD as compensation for their time.

**Table1.** Characteristics of participants (N = 41)

| Table 1. Characteristics of participants (N = 41) |  |  |
| --- | --- | --- |
| Variable | Median | Range |
| Age (years) | 40 | 25-65 |
|  | Percent | Frequency |
| Gender |  |  |
| *Male* | 51% | 21 |
| *Female* | 49% | 20 |
| Ethnicity |  |  |
| *French* | 15% | 6 |
| *Caucasian* | 12% | 5 |
| *Carribean* | 12% | 5 |
| *East and Southeast Asian* | 12% | 5 |
| *Southern European* | 10% | 4 |
| *North American Indigenous* | 7% | 3 |
| *Black and African* | 7% | 3 |
| *South Asian* | 7% | 3 |
| *Mixed* | 7% | 3 |
| *Northern European* | 5% | 2 |
| *Eastern European* | 2% | 1 |
| *Other North American* | 2% | 1 |
| Marital Status |  |  |
| *Married/common-law* | 71% | 29 |
| *Single* | 19% | 8 |
| *Divorced/widowed/separated* | 10% | 4 |
| Income |  |  |
| ≤ \$29,999 | 5% | 2 |
| \$30,000 - \$79,999 | 53% | 22 |
| ≥ \$80,000 | 42% | 17 |
| Level of education completed |  |  |
| *High School* | 24% | 10 |
| *College* | 42% | 17 |
| *University* | 29% | 12 |
| *Post Graduate* | 2% | 1 |

**Table2.** Characteristics of participants by focus group

| Table 2. Characteristics of participants by focus group |  |  |  |  |  |  |
| --- | --- | --- | --- | --- | --- | --- |
|  | Sudbury 1 | Sudbury 2 | Sudbury 3 | Missis-<br>sauga 4 | Missis-<br>sauga 5 | Missis-<br>sauga 6 |
| Number of participants | 8 | 6 | 6 | 7 | 7 | 7 |
| Median age in years (range) | 48 (35-62) | 33 (27-35) | 48.5 (39-65) | 55 (35-59) | 30 (25-33) | 44 (36-63) |
| Gender |  |  |  |  |  |  |
| Male | 4 (50%) | 3 (50%) | 3 (50%) | 4 (57%) | 3 (43%) | 4 (57%) |
| Female | 4 (50%) | 3 (50%) | 3 (50%) | 3 (43%) | 4 (57%) | 3 (43%) |
| Ethnicity |  |  |  |  |  |  |
| French | 2 (25%) | 1 (16.7%) | 3 (50%) | - | - | - |
| Caucasian | 1 (12.5%) | - | - | 1(14.2%) | 1(14.2%) | 2(28.5%) |
| Caribbean | - | - | - | 1(14.2%) | 2(28.5%) | 2(28.5%) |
| E and SE Asian | 1 (12.5%) | 1 (16.7%) | - | - | 1(14.2%) | 2(28.5%) |
| S European | - | 1 (16.7%) | 1 (16.7%) | - | 1(14.2%) | 1(14.2%) |
| NA Indigenous | 2 (25%) | 1 (16.7%) | - | - | - | - |
| Black/African | - | 1 (16.7%) | - | 2(28.5%) | - | - |
| South Asian | - | - | - | 2(28.5%) | 1(14.2%) | - |
| Mixed | - | 1 (16.7%) | 1 (16.7%) | - | 1(14.2%) | - |
| N. European | 1 (12.5%) | - | 1 (16.7%) | - | - | - |
| E. European | 1 (12.5%) | - | - | - | - | - |
| Other N. Am. | - | - | - | 1(14.2%) | - | - |
| Marital Status |  |  |  |  |  |  |
| Married/c. law | 6 (75%) | 5 (83.3%) | 6 (100%) | 5(71.4%) | 2 (28.6%) | 5(71.4%) |
| Single | 2 (25%) | - | - | 1(14.3%) | 5 (71.4%) | - |
| Div./wid./sep. | - | 1 (16.7%) | - | 1(14.3%) | - | 2(28.6%) |
| Income |  |  |  |  |  |  |
| ≤ 29,999 | 1 (12.5%) | - | - | 1(14.3%) | - | - |
| 30,000 - 79,999 | 7 (87.5%) | 2 (33.3%) | - | 6(85.7%) | 5 (71.4%) | - |
| ≥ 80,000 | - | 4 (66.7%) | 6 (100%) | - | 2 (28.6%) | 7 (100%) |
| Education |  |  |  |  |  |  |
| High School | 3 (37.5%) | 1 (16.7%) | 2 (33.3%) | 2(28.6%) | - | - |
| College | 5 (62.5%) | 3 (50%) | 2 (33.3%) | 2(28.6%) | 4 (57.1%) | 3(42.9%) |
| University | - | 2 (33.3%) | 2 (33.3%) | 3(42.9%) | 2 (28.6%) | 4(57.1%) |
| Post Graduate | - | - | - | - | 1 (14.3%) | - |

### Patient and Public Involvement

The central research question - how do members of the general public perceive the use of health data for AI research - was directly informed by the results of previous qualitative studies with 60+ members of the public. Before the research was started, the draft scenarios were reviewed and refined based on feedback from the Manager of Public Engagement at ICES and multiple members of the public, including students at the University of Toronto and friends and family members of Vector staff. The corresponding author, PAP, is co-author of the Consensus Statement on Public Involvement and Engagement with Data-Intensive Health Research and the Lead for the Public Engagement Working Group of Health Data Research Network Canada. Through those and other initiatives, PAP has connections to multiple patient and public advisors, from whom the research team will seek advice when disseminating study findings, including through non-academic channels such as “The Conversation” and social media.

### Data Collection

Focus groups were moderated by an experienced male focus group moderator employed by Edelman (10 years of professional experience, RIVA-trained) with no prior relationship with the participants. The moderator was hired to conduct the focus groups. He had no prior knowledge about AI/ML and had no vested interest in the outcome of this project. This information was disclosed to participants at the beginning of the session. Having an external facilitator enabled the research team to benefit from the experience of a skilled professional, provided an environment in which participants would be more likely to feel free to express negative opinions about AI and the Vector Institute than if a member of the Vector Institute staff were facilitating, and allowed the research team to focus on observing the participant discussion and taking field notes. The discussions followed a semi-structured discussion guide (see supplemental material) which allowed for free-flowing conversation as well as facilitated discussion of written scenarios, with prompts on certain questions. All members of the research team (MM, TS, PAP) observed every focus group from behind a one-way mirror and took independent field notes during the sessions. Focus group participants were informed that researchers were in attendance behind the one-way mirror, and that sessions were audio-recorded. Audio-recordings were transcribed verbatim by Edelman and participant names were replaced with a code (e.g., M01 for male 1) before the transcripts were provided to the research team for analysis.

### Data Analysis

Data were analysed by MDM, TS and PAP using a qualitative descriptive approach which is a naturalistic form of inquiry that aims to remain “data-near” while inductively interpreting and thematically grouping and detailing respondent experiences, beliefs and expectations[27–28]. MDM, TS and PAP worked together to develop the descriptive coding framework based on the verbatim transcripts and field notes taken during the focus group sessions. The transcripts were read and re-read as coding was performed independently by MDM and TS using a combination of Microsoft Word and Microsoft Excel software. No software was used to supplement human qualitative coding. MDM, TS and PAP used an inductive analytic approach to derive themes based on the data and socialised and refined themes through group discussion. Differences in opinion between MDM, TS and PAP were resolved through iterative discussions. Review and coding of transcripts stopped when inductive thematic saturation was achieved, i.e., when MDM, TS and PAP agreed that additional coding and thematic analysis would not result in any new codes or themes. Though the sample was not designed or intended to provide information about variation in perspectives based on gender, location or age, the research team analysed the theme-coded statements for each of those characteristics and did not find any consistent or significant correlations. The research team was open to the possibility of recruiting additional participants for additional focus groups if there was insufficient data to identify themes; however, based on the finding that themes were strong and consistent across the focus groups, no additional participants were recruited. No formal participant feedback was sought, although the interviewer continually reflected focus group participants’ views back to participants to ensure that their views were being captured adequately.

## RESULTS

The analysis identified mixed, mostly negative views about AI in general. There were three major themes from the participants discussion of the realistic health AI scenarios, (i) participants had hopes for health AI and perceived benefits from it, (ii) they also identified AI-specific concerns and fears and (iii) they described the conditions under which they supported the use of health data for AI research. Finally, though it was not an objective of the study, the realistic health AI scenarios were found to have an educational effect.

### Theme 1: Mixed, mostly negative views about AI in general

Participants had mixed views about AI, but mostly unfavourable perceptions (Box 1). Negative comments referred to the potential for job loss, lack of human touch, and humans losing control over AI, with multiple references to malicious robots (e.g., Terminator, HAL 9000). Several participants shared stories of advertisements being presented to them on their mobile phones after they had spoken about a topic, which they interpreted as proof of AI surveillance of their behaviour. Some participants expressed hope for AI in terms of autonomous vehicles, AI’s perceived ability to increase convenience and the ways that AI could be useful in dangerous environments not suitable for humans. However, most of the participants who expressed positive statements about AI also noted concerns given uncertainty about how AI will affect society.

##### Box 1: Mixed, mostly negative, views about AI in general

1. I feel like it’s one of those things that we’d all be diving headfirst towards, but may be something that could have long-term implications for us as a society down the road that maybe we didn’t fully understand when we dove into it at first. (M015-Mississauga2)
2. So, when I think of AI, I have mixed feelings about it because I think about, "Will my job exist in the future, or will most jobs exist in the future?" …. I think very few of us actually know what AI could be in the next year, ten years, 50 years from now. (F017-Mississauga2)
3. Are we phasing ourselves out? (M008-Sudbury3)
4. I think it’s impersonal. Not like that human touch. Where there’s substance and feelings or emotions. (F002-Sudbury1)
5. It’s portrayed as friendly and helpful, but it’s always watching and listening… So I’m excited about the possibilities, but concerned about the implications and reaching into personal privacy (M007-Sudbury2)
6. You talk to somebody about something and then an ad will pop up on your phone for it. It’s almost like you’re being listened to (F008-Sudbury3)
7. Scary. Out of control… are they [AI] going to take over. It’s going to be jobless. (F004-Sudbury1)

### Theme 2: Hopes and perceived benefits of health AI

Participants perceived benefits from each of the three realistic health AI scenarios (Box 2). Perceived benefits were both epistemic (e.g., the perception that AI could generate knowledge that would otherwise be inaccessible to humans) and practical (e.g., the ability of AI to sift through data, perform real-time analyses and provide recommendations to health care providers and directly to patients). Of the three scenarios presented (Table 3) participants saw the greatest benefit of the AI-based Cancer Genetics Test, where it was perceived that AI could save lives. Participants also commented favourably on the benefits of an AI-based app for older adults helping people maintain independence and about the potential for a large laboratory test results dataset to support health AI training, education and discovery research (Table 3).

**Table 3.** Summary of main participant views on three realistic health AI research scenarios

| Health AI research Scenario | Main Hopes and Perceived Benefits | Main Fears and Perceived Risks | Main Conditions for Scenario to be Acceptable |
| --- | --- | --- | --- |
| AI-based Cancer Genetics Test:<br>Academic researchers applying ML to consented genetic data to study cancer cell evolution | Potential for AI to save lives by identifying origin of cancers so treatment can be tailored<br>AI provides faster and more accurate results than would be possible with humans<br>AI has capability to analyse more data than humans could | Risk of re-identification because genetic material can never be truly anonymous<br>Concerns related to spread of AI application outside of beneficial cancer scenario (e.g., misuse of AI tool for inappropriate prenatal genetic screening) | AI must be used as a tool with a human (doctor) making the final decision<br>Data must not be sold (reference to 23andMe partnership with Glaxo Smith Klein)<br>Participants noted and responded positively to the fact that data subjects in the fictional scenario had provided consent for data to be used for AI research |
| AI-based App to Help Older Adults Aging at Home: Team of academic and industry researchers using ML to develop a mobile phone application (app) to help older adults self-manage chronic conditions and age at home | AI creates a useful tool that provides helpful information to patients<br>AI helps address health human resource shortages<br>AI is helpful for people who do not have family and friends to support them | Concern that machines and AI will inappropriately be viewed as a substitute for human interaction | AI-based app supplements versus replaces human care<br>People using the AI app would need to be fully aware that it is capturing and using their data (transparency)<br>People have the option/choice to not use the AI app |
| Accessible Dataset with Lab Test Results for AI: Creation of a large accessible de-identified dataset of unconsented laboratory test results to be used a foundation for multiple AI-related purposes | Ability to use AI to generate new knowledge from large amounts of data<br>AI analysis faster and more efficient than humanly possible<br>Utility of dataset for teaching AI | Absence of specific purpose or intended benefit from AI<br>Concern about misuse when companies access health data | External organisation certifies that data are de-identified<br>Some participants would only support scenario if data subjects provide consent |

##### Box 2: Hopes and perceived benefits of health AI

1. There’s just so much potential value… this can potentially save lives. (M017-Mississauga2)
2. It could be a help worldwide to see similar symptoms…it will be quicker because using AI in a computer, you’ll be able to get that data and those analytics quicker. (F003-Sudbury1)
3. I think it’s fantastic. The more data they collect, the more they’ll be able to identify the patterns of these cancers and where they originate from. I think it’s just great. (F009-Sudbury3)
4. A lot of times doctors are very busy… So if they have a database or something where they could put in a particular disease or something they’re suspecting, and then this database just brings up - narrows down what the possibilities are. That might be better. (F013-Mississauga1)
5. If I could do that as an elderly person and keep my integrity and pride and myself, like staying home instead of having to be placed in a long term care facility. And this little [AI-based] app can help me to stay home and not have a nurse come in my house two, three times a day. (F002-Sudbury1)
6. When you can reach out and have a sample size of a group of ten million people and to be able to extract data from that, you can’t do that with the human brain. A group, a team of researchers can’t do that. You need AI. (M018-Mississauga3)
7. You put everything into a data[set], somebody’s going to learn something on that. (M002-Sudbury 1)

### Theme 3: Fears and perceived drawbacks of health AI

Participants were primarily concerned that the health data provided for one health AI purpose might be sold or used for other purposes that they do not agree with (Box 3). They also expressed AI-specific concern about the lack of human touch when machines are deeply integrated into care, potential job losses and the potential for AI to lead to a decrease in human skills over time if people become “lazy” and overly reliant on computers. Some additional fears and concerns specific to the individual scenarios were noted including, inability to guarantee privacy when genetic information is used for AI, skepticism that older adults would be able to use an AI-based app, and concern about companies misusing or selling data.

##### Box 3: Fears and perceived drawbacks of health AI

1. There’s no guarantee that they [the people developing AI] are going to have any kind of integrity or confidentiality or anything like that. (F003-Sudbury1)
2. Are they going to take my information, are they going to sell it? So, it kind of makes you scared when other companies are buying it. (F016-Mississauga2)
3. For me the big question is ownership of that data. (M018-Mississauga3)
4. I don’t find it very appropriate. First of all, it’s going to take jobs away from health professionals. If the app has to tell them, suggest things or whatever, there’s no communication there, like face-to-face. (F010-Sudbury3)
5. But it also misses out on that human component where the [personal support worker] comes in and talks to you and things like that. (M007-Sudbury2)
6. The concern is always that you lose some of those soft skills. And how many times in the medical field have you heard that a nurse practitioner or a doctor went on a hunch and found out what the problem was. So that’s a concern, that you lose some of those soft skills and that relies on intuition when you rely solely on AI, on computers and programs and algorithms. (M010-Sudbury3)

### Theme 4: Conditions under which health AI scenarios are more acceptable

Many participants suggested specific conditions that would make health AI acceptable to them, the most common requirement was that AI be used as a tool that helps humans make decisions versus an autonomous decision-making system (Box 4). In addition, across multiple scenarios, participants stated the requirement for transparency about how data are used in health AI, often expressed in terms of their preference that data subjects be fully informed about how data will be used and given the option of providing informed consent or opting out.

##### Box 4: Conditions under which Health AI Scenarios are More Acceptable

1. As long as it’s a tool, like the doctor uses the tool and the doctor makes the call. As long as the doctor is making the call, and it’s not a computer telling the doctor what to do. (M001-Sudbury1)
2. But I think that it should be stressed for the people that are going to be using it, that it should not be their primary source of health information. They shouldn’t skip going to the doctors. This is to be used in conjunction with that. (F007-Sudbury2)
3. The data may be used for research, but they may not be fully aware of it. They may have clicked "I accept" and that part was like - I was like, "That’s kind of tricky, kind of." (F002-Sudbury1)
4. That’s the thing that threw me off… it was the fact that you didn’t get to choose that your information gets used in this process… "Give me a choice." (M012-Mississauga1)
5. Transparency…Why are they even taking the data in the first place? How would it help people in the future? Just understanding the purpose behind all of this. (M017-Mississauga2)

### Theme 5: Educational effect of realistic health AI scenarios

There was a significant difference between the dystopian and/or utopian statements of participants at the beginning of each focus group (Box 1) and their comments about the health AI scenarios (Boxes 2, 3, 4, 5 and Table 3) which tended to be more grounded in reality. In some cases, participants were direct in stating that the health AI scenarios had an educational effect for them (Box 5).

##### Box 5: Educational Effect of Realistic Health AI Scenarios

1. I think our discussion prior to any of these scenarios was more geared toward just generally based [AI], wasn’t more toward the health… I didn’t think it was so appropriate but then seeing the other two [health AI] scenarios with it [the third AI scenario], I think it could all go hand in hand in the healthcare system. I’m leaning more towards it than my opinion was before. (F006-Sudbury2)
2. I’m not usually that positive, but I’m pretty positive about all of it, everything that we read [the health AI scenarios] so far… I’m anti-computer… But everything I’ve seen so far… I think it’s all good information and it’s all good tools, but the keyword "tool." It’s a tool. And I see this being an awesome tool as well. (M004-Sudbury1)
3. [Before Scenarios] You can create a Terminator, literally, something that’s artificially intelligent, or the Matrix… it goes awry, it tries to take over the world and humans got to fight this. Or it can go in the absolute opposite where it helps… androids… implants… Like I said, it’s unlimited to go either way. [After reviewing health AI scenario] I know what they’re trying to get done. I agree with all these things. I think they’re extremely beneficial for everyone… So now I can say, you know what, I’m confident that this is going in the direction of where I would like this to go because I can’t find a downside to an app like this. (M020-Mississauga3)

## DISCUSSION

Upon reflecting and discussing the health AI scenarios, participants demonstrated mixed, but generally positive views of the application of AI in health research, provided certain risks were mitigated and conditions were met. Consistent with the literature, this study found that members of the general public have little understanding of AI and ML in general. Given this low level of knowledge, dystopian and utopian extremes presented in the media, and uncertainty about the future of AI and ML which runs across society, the term “hopes and fears” is likely a better fit than “benefits and risks” to describe how members of society perceive AI[15,16].

Overall, participants’ perception of three realistic health AI scenarios were more positive than their perception of AI in general. Many of the views expressed by participants were similar to the findings from a systematic review of public views of data-intensive health research[10] which found general support for using of health data for research with some conditions, concerns about privacy and security, the requirement that there be a public benefit, more trust in public sector studies compared to private sector studies, and varying views on the need for consent. This study adds information about participants’ AI-specific hopes (e.g., potential for faster and more accurate analyses, ability to use more data), fears (e.g., concern that AI will be used for objectionable purposes, lack of human touch, decrease in human skills over time due to over-reliance on machines) and conditions for acceptability (e.g., a human must be in the loop for computer-aided decisions).

Consistent with previous studies of public perspectives about health AI[16,23–25], participants’ support for health AI scenarios was linked to their perceived public benefit of the scenarios, with people being most supportive when they believed that AI could bring an important new capability to a problem beyond what humans could contribute. Each of three health AI research scenarios were viewed as being acceptable by most of the participants of the focus groups (Table 3). Of the three scenarios, the AI-based Cancer Genetics Test was the most supported, with several participants linking their support to personal or family experiences with cancer. The next highest supported scenario was the AI-based App to Help Older Adults Aging at Home. Participants were generally supportive of the scenario focused on creating a large accessible dataset, but were direct in stating that the benefits from it were less clear to them. Though care was taken to construct scenarios focused on health AI research, participants’ support was mostly associated with the benefit expected from the final health AI application, even when scenarios highlighted the fact that there was no guarantee that the research would achieve its intended impact. Given the Gartner Hype Cycle,[29] this may present a risk for AI/ML research. If members of the public assume that health AI research will always be successful, there is increased likelihood of disillusionment, potentially leading to an AI winter and decrease in research funding for AI/ML.

In this study, many participants’ concerns with the health AI scenarios were not directly related to AI. As has been observed for data-intensive health research in general, people were concerned about lack of transparency, and potential abuses and misuses of their health data, particularly when companies work with health data[10,13]. High profile news stories about data breaches as well as coverage of lawsuits (e.g., related to Google[20,21]) can heighten these concerns. In addition, participants did have some fears and concerns related to health AI which were very similar to the concerns that they expressed about AI in general, e.g., fear that AI would take over decisions and/o result in job losses.

Consistent with the available small literature on public views about health AI,[15–19, 23–25] the main condition for the health AI scenarios to be seen as acceptable was that AI be used as a tool by humans, and that humans continue to be in the loop. This condition is not surprising given the general fears associated with all AI, and also aligned with the American Academy of Dermatology Position Statement on Augmented Intelligence (their preferred term over artificial intelligence) which refers to “symbiotic and synergistic roles of augmented intelligence and human judgment”[30]. Participants’ support was also conditional on transparency about how data are used for health AI. Some were direct in stating that consent should be obtained before data are used for health AI, while other participants noted that current consent processes (e.g., long forms) are not the solution, and many emphasised the need for plain language explanations of how data are used for health AI, preferably delivered by a human. Again, this finding is aligned with the American Academy of Dermatology Position Statement which states “there should be transparency and choice on how their medical information is gathered, utilised, and stored and when, what, and how augmented intelligence technologies are utilised in their care process.” In this regard, the views of the general public about health AI are similar to their views on data-intensive health research in general;[10,11] i.e., they have mixed views on consent with most people primarily wanting to know if, how and when their data were used for research.

Taken as a whole, the findings of this study and other qualitative research should influence health AI research and application. Given widespread uncertainty about exactly how AI will impact society, and increasing use of public data (including unconsented data) for AI, we need to understand which uses of health data for AI research are supported by the public, and which are not. Transparency and plain language communication about health AI research are necessary but not sufficient[31]. This is not simply a matter of informing members of the public about how health data are used in AI research. Consistent with the Montreal Declaration for Responsible Development of AI[32] the objective should be to take the science of health AI in directions that the public supports. By behaving in a trustworthy manner, respecting public concerns and involving members of the public in decisions related to health AI, we can align with the Consensus Statement on Public Involvement and Engagement with Data-Intensive Health Research[33] to establish socially beneficial ways of using health data in AI research.

### Limitations

This study has limitations. Foremost, results may not be generalisable across or outside of Ontario. It is possible that participants from other settings, e.g., rural Ontario, remote northern Ontario, specific sub-populations or other jurisdictions would have different views. Given the low level of knowledge about AI in general it is possible that the views of participants would change substantially if they learned and understood more about AI. There are many uses of health data for AI which were not included in the scenarios in this study, and it is possible that participants would have different views if the scenarios were different or altered.

## Data Availability

All data relevant to the study are included in the article. All authors had access to all the data in the study and take responsibility for the integrity of the data and the accuracy of the data analysis. No unpublished data are available outside of the study team.

## FUNDING STATEMENT

This research was funded by the Vector Institute.

## COMPETING INTERESTS STATEMENT

MDM has nothing to disclose.

TS has nothing to disclose.

PAP has nothing to disclose.

## AUTHORSHIP STATEMENT

All authors contributed to the design of the study, attended all focus groups, developed and refined the themes, contributed text directly to the manuscript, and approved the final version submitted for publication. Melissa McCradden and Alison Paprica led the literature review. Alison Paprica led the work to design the scenarios with contributions from Tasmie Sarker, Melissa McCradden and multiple other individuals who are acknowledged. Tasmie Sarker and Melissa McCradden both independently coded all transcripts. Alison Paprica reviewed all coding and performed analyses with Melissa McCradden and Tasmie Sarker to develop the descriptive coding framework and identify themes. Alison Paprica was the lead for preparation of the manuscript. All of the authors gave approval of the final version for publication and agreed to be accountable for all aspects of the work.

## ACKNOWLEDGEMENTS

The authors thank Elham Dolatabadi, James Fernandez, Ian Gormely, Jenine Paul, Céline Moore, Andrea Smith, and Linda Sundermann for their contributions to the health AI scenarios.

## Notes

### Competing Interest Statement

The authors have declared no competing interest.

